# Animal Model for Pericardium Patch Thoracoabdominal Aortic Aneurysm: Step by Step

**DOI:** 10.1101/2024.10.20.24315711

**Authors:** Allana Tobita, Bruno Jeronimo Ponte, Maria Fernanda Cassino Portugal, Anna Paula Weinhardt Baptista, Igor Rafael Sincos, Nelson Wolosker

## Abstract

2.

**Objectives:** We aimed to improve previously described methods for confection of thoracoabdominal aortic aneurysms (TAAA) in porcine animal models, reducing surgical procedure time and specimen mortality.

**Methods:** A total of 18 swine underwent a surgical procedure in order to confect a TAAA. All anesthetic precautions were taken. All the procedures were carried out via laparotomy and retroperitoneal access. An autologous peritoneum patch was used to make the aneurysm in 2 animals, and a bovine pericardium patch in 16. All the animals were followed up post-operatively, and were re-approached after 4 weeks for analysis of the aneurysm sac. The models that did not die in the post-operative period were euthanized under institutional recommendations

**Results:** All the animals underwent laparotomy with retroperitoneal access. Two were submitted to an autologous peritoneum patch and 16 to a bovine pericardium patch. Three models underwent single suprarenal clamping; while 15 underwent sequential clamping. There were no differences in surgical time *(p 0.207)* or total clamping time *(p 0.276)* between groups. There was a higher mortality rate after 4 weeks in models submitted to single clamping (100%) compared to sequential clamping (26.7%) *(p 0.0017)*.

**Conclusion:** The experimental model of a thoracoabdominal bovine pericardium aneurysm which uses a sequential clamping technique, provides a stable and reliable platform with consistent anatomy and patency for up to four weeks. This model can be extremely valuable for assessing new endovascular therapy options in living organisms

## 3. Introduction

Over the years, experimental models have been crucial in deepening our understanding of the pathophysiology of various diseases and have played a major role in the search for effective therapeutic interventions. Animal experimental trials are particularly important in fields where applicable *in vitro* models are limited. Although these studies are subject to strict legal and ethical restrictions, they are essential for developing safe protocols that can be directly applied to human patients. They represent an important stage in evaluating the efficacy of new medical devices and pharmacological therapy before their application in clinical trials.(1,2)

Despite the progress in surgical techniques, diagnosing and treating pathologies of the thoracoabdominal segment of the aorta remains challenging. Experimental models are crucial in evaluating disease evolution and testing new therapies in this field. (3–9)

Several groups have described the development of animal models for studying thoracoabdominal pathologies with significant variation. (10) However, surgical models have been limited in invasiveness, and establishing a proper intensive care system to support the high morbidity and mortality associated with procedures of this magnitude is challenging.

Our experience with experimental porcine models has helped us develop procedures to improve animal handling and reduce complications, especially when working with swine in thoracoabdominal intervention. (8,9) We have recently created a protocol to study the hemodynamic effects of the Multilayer Flow Modulator Stent (MFMS - Cardiatis, Isnes, Belgium) in artificially induced thoracoabdominal aneurysms in swine models.(9) This latest study represents the culmination of our work and serves as a refinement of practices that led to developing these procedural standards.

This report outlines our procedures to assist other researchers in developing protocols for implementing an experimental animal model for thoracoabdominal aneurysms with minimal complications and animal loss.

## 4. Methods

The study was conducted between November 2016 and April 2019 at the Center for Experimentation and Training in Surgery of a quaternary hospital in São Paulo, Brazil. This centre is accredited by the Association for Assessment and Accreditation of Laboratory Animal Care and Use Committee. The completed Animal Research Reporting In Vivo Experiments guidelines checklist is in the Appendix.

### 4.1. Animal selection and preoperative anaesthetic procedures

Eighteen Large white pigs, aged 4 to 10 months and weighing 37 to 75 kg, were assigned.

Before surgery, all pigs were instructed to fast, refraining from consuming food or drinks for 12 hours. The protocol required pre-anesthesia with ketamine (10.0 mg/kg) and midazolam (0.25 mg/kg) administered intramuscularly.

A 22-gauge BD Insyte catheter (BD Infusion Therapy Systems Inc, Sandy, Utah) was used to catheterize the marginal ear vein for venous access. The right carotid artery was catheterized to measure invasive arterial pressure.

To induce anesthesia, the pigs were given etomidate (1 mg/kg) and propofol (5 mg/kg). Size 7.0 Portex endotracheal tubes (Smiths Medical, Ashford, UK) were used for intubation. 1.5% isoflurane was used for inhalational anesthesia, with the ventilator set at a tidal volume of 10 mL/kg. Fentanyl (2.5 mg/kg) maintained anesthesia.

Fluid replacement was carried out using a maintenance crystalloid solution at a rate of 10 mL/kg/h. Additionally, a crystalloid solution at a rate of 1 to 2 mL/kg/h was given as needed in bolus form to maintain a mean blood pressure of at least 70 mm Hg. The protocol indicated that animals with unresponsive hypotension should be excluded from the study.

All animals received antibiotic prophylaxis with penicillin G benzathine (2.4 million units IM) and cephazolin (1 g IV).

### 4.2. Intraoperative Technique

The initial protocol reproduced Maynar’s technique, where the aneurysmal sac was created using the native peritoneum. (10) The aneurysmal patch was made of peritoneal tissue during the surgery in the first two swine models. However, this technique prolonged the operation and had hemodynamic consequences, so it was abandoned for the third and following animals.

### 4.3. Aneurysmal patch creation

In sixteen cases, the aneurysmal patches were prepared in a back table before the start of the procedure. A bovine pericardium patch (Braile Biomedica, São José do Rio Preto, São Paulo) was folded in half, and a trapezoid pattern was drawn on the patch sheet with a surgical marker *(Figure 1).* Then, that sheet was molded into two equal trapezoid leaflets and sutured into an oval configuration. A continuous suture of Prolene 5-0 was used. The approximate final dimensions of the patch were 7 cm in length and 4,5cm in width.

**Figure 1:**
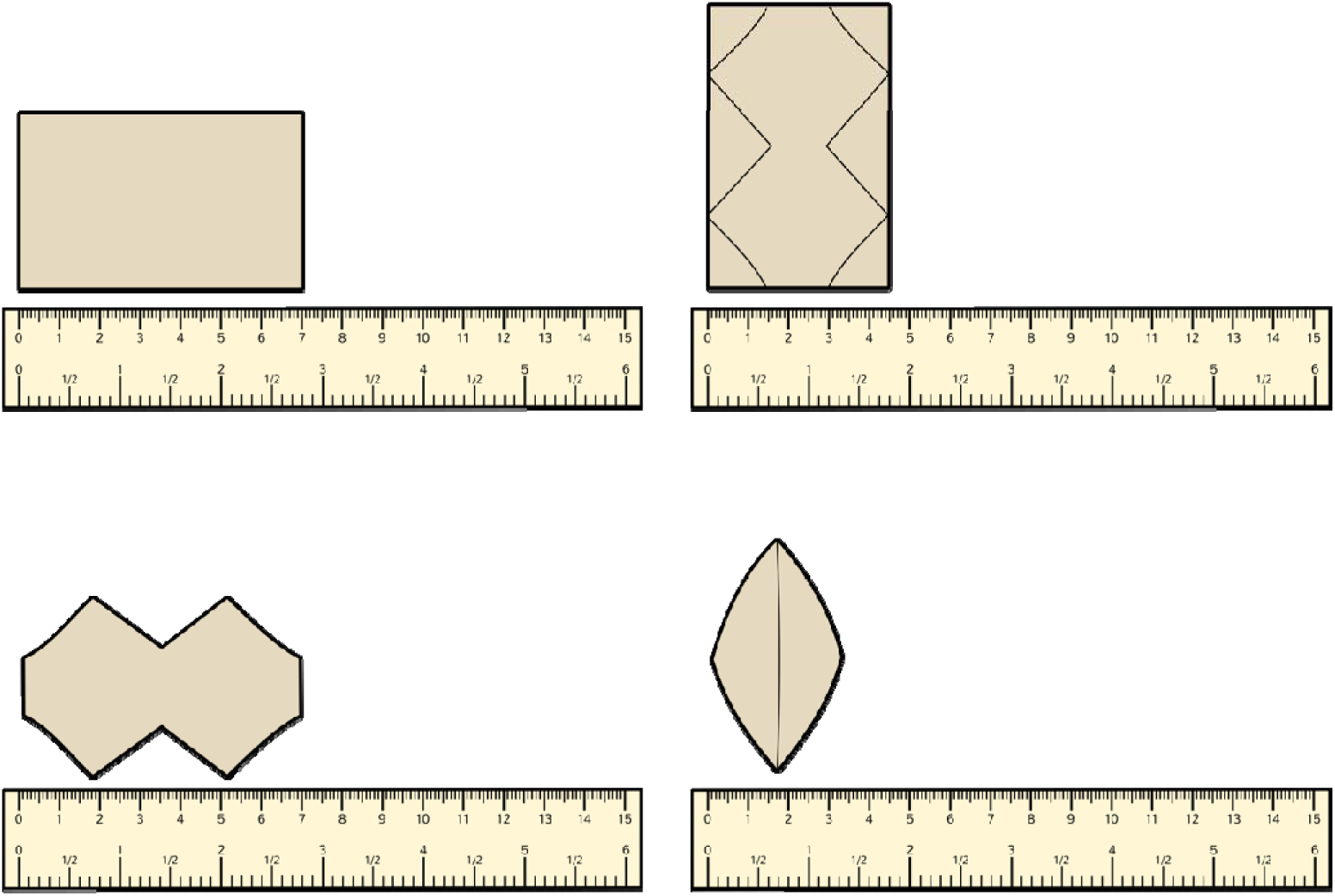
**A:** The bovine pericardium patch; **B:** The trapezoid shape being drawn on the pericardial patch; **C**: Two trapezoid-shaped leaflets were made; **D**: The trapezoid-shaped pericardium was molded into an oval configuration after a continuous suture using Prolene 5-0.

### 4.4. Surgical access and aneurysm creation

All animals underwent laparotomy with retroperitoneal dissection to access the visceral aorta, that was controlled at the top (at the level of superior mesenteric artery) and bottom (at the iliac bifurcation level). The renal and lumbar arteries were also repaired during the procedure.

Additionally, all subjects received systemic heparin dose of 200 UI/kg two minutes before the proximal and distal clamping of the aorta.

### 4.5. Clamping Type and Intraoperative Technique

**Single clamping**: In the first three swine models, the aorta was clamped to the renal arteries simultaneously. This technique led to a longer suprarenal clamping than the sequential clamping and significantly impaired intraoperative hemodynamic balance. As a result, this approach was abandoned from the fourth animal onwards.

**Two-step sequential clamping**: In 15 animals, a clamp was placed on the aorta just below the renal arteries. Once the clamp was secure, a 6 cm long incision was made just below the emergency of the renal arteries, and a thin elliptic patch of the aortic wall measuring 3 mm was removed. The aneurysmal patch, composed of bovine pericardium, was then sutured to the incision using a continuous suture of Prolene 4-0.

After that, the clamp was moved 1 cm upward along the aorta (second step), above the renal artery ostium. The incision was then extended, and the proximal part of the patch was sutured to the aorta at the level of the renal arteries, thus completing the creation of the aneurysmal sac. The two-step sequential clamping is illustrated in Figure 2, and the final aspect of the saccular aneurysm in Figure 3.

**Figure 2-.**
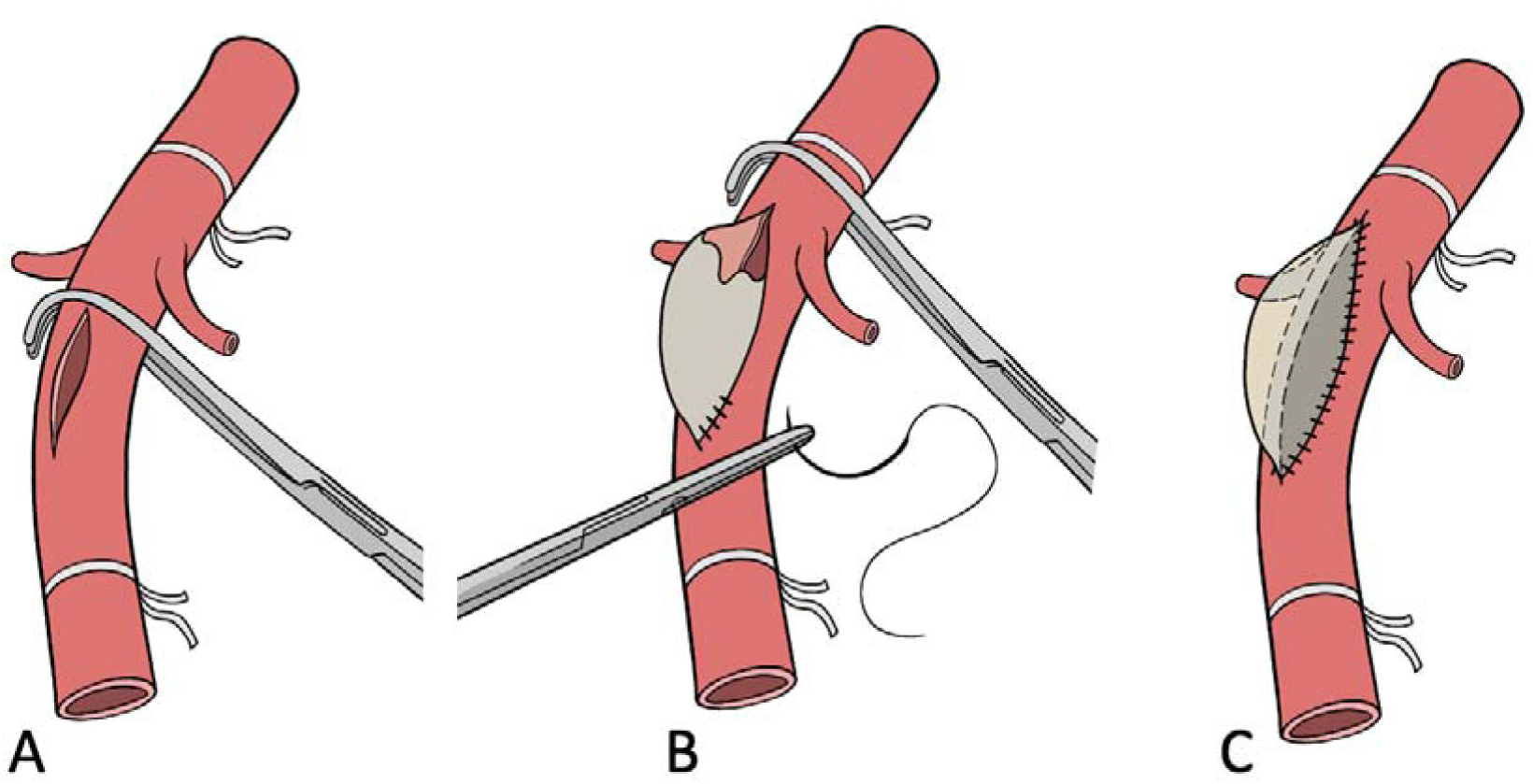
Two-Step Sequential Clamping. Initially, the clamp is placed under the renal artery **(A)**. A 6cm incision is made and the distal part of the patch is sutured at the aortic wall. After that, the clamp is moved 1cm above the renal artery **(B)**, completing the suture and the creation of the aneurismal sac is concluded **(C)**.

**Figure 3-.**
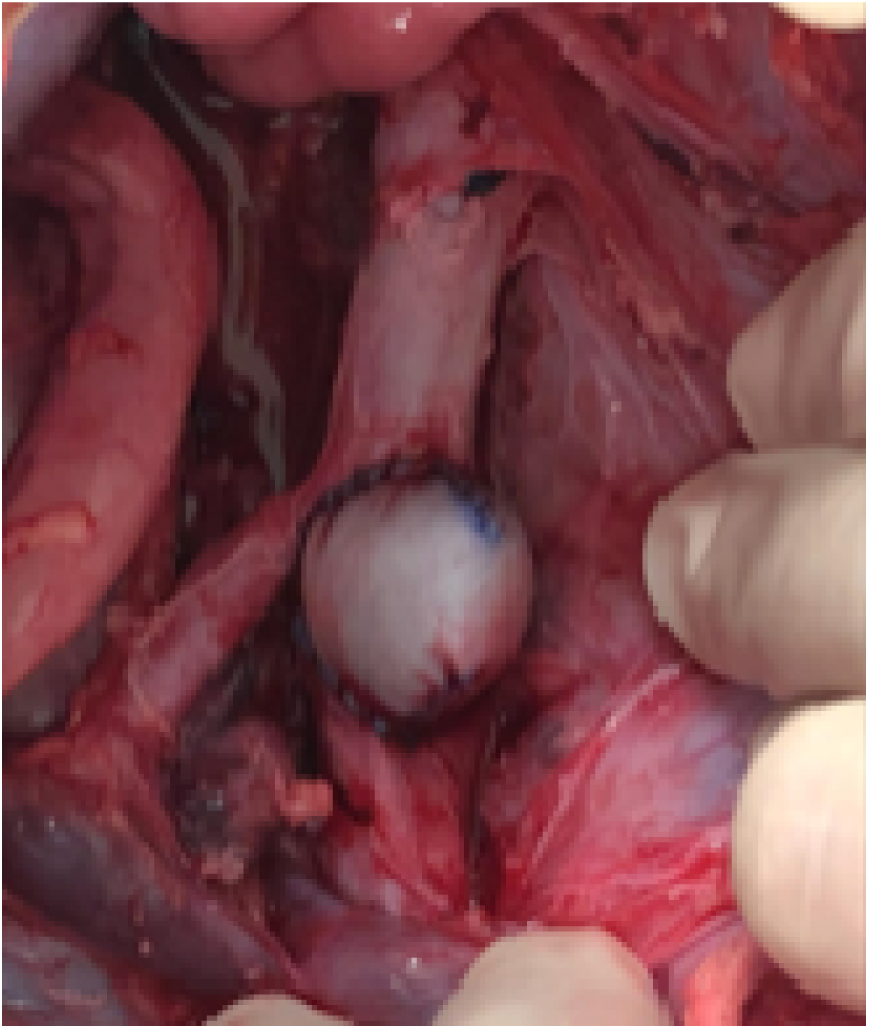
Final aspect of the saccular aneurysm.

After careful hemostatic revision, the aortic flow was restored, and the renal and lumbar arteries were unclamped.

### 4.6. Hemodynamic and Electrolytes Assessment

Blood samples were collected before aortic clamping and whenever needed during the procedure for monitoring of hematic and electrolytic patterns using the portable analyser i-STAT (Abbott).

### 4.7. Intraoperative imaging control and technical success assessment

After restoring blood flow, we performed Intraoperative aortography using a 5F *PigTail* catheter (Impulse^TM^, Boston Scientific) with radiopaque centimeter markings positioned at the level of the first lumber vertebrae and inserted through the femoral artery. We measured the diameter and extent of the aneurysm and the healthy inframesenteric aorta (Figure 4).

**Figure 4-.**
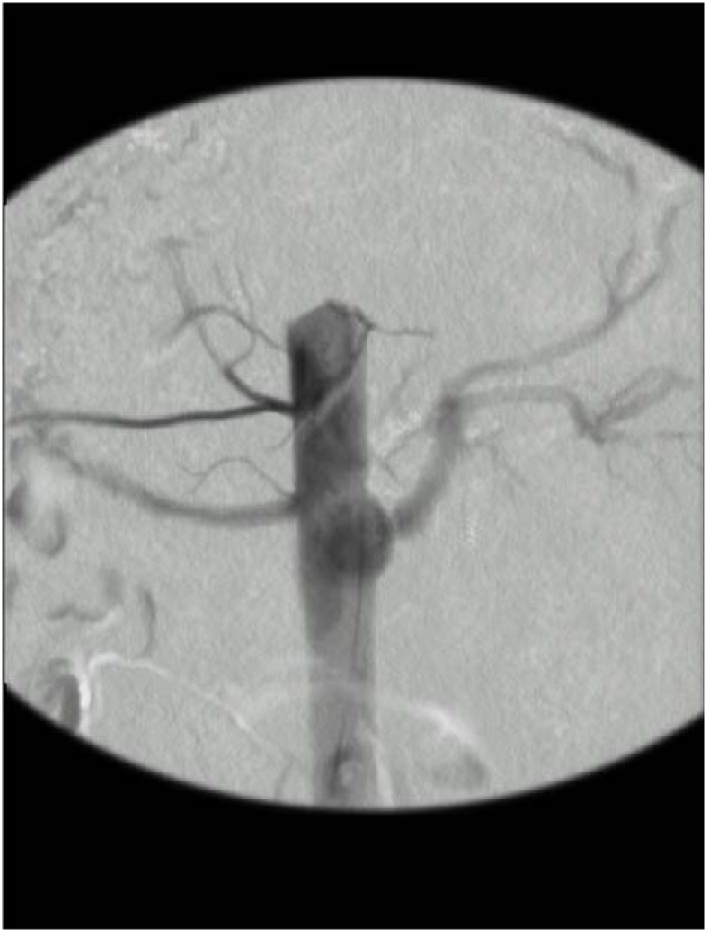
Final aspect after aortography, demonstrating the creation of a saccular aneurysm.

We considered technical successes as a case in which the aneurysmal section showed an enlargement of at least 50% of the healthy diameter of the aorta.

### 4.8. Post-operative protocol

To prevent a slowdown of the intestines, animals were refrained from eating solids for 24-48 hours after surgery. Water was provided freely after 24h. When the animals started eating again, they were given a dog ration (Hill’s Critical Care © 2018 Hill’s Pet Nutrition, Inc) until they fully accepted their normal dry diet.

Fluid therapy began on the day before surgery with a solution containing 1500mL + NaCl 0,9% 1000mL + 50% glucose 100mL every 8h, given every 8 hours for the first 48 hours and adjusted as needed.

Following veterinary determinations, pigs were given daily gastric protection medication (Ranitidine2mg/Kg, by intravenous or intramuscular injection, thrice daily), and when necessary, an antiemetic (Metoclopramide 0,5mg/Kg, by intravenous or intramuscular injection up to thrice daily), and pain relief (Ketoprofen 5mL by intravenous injection, once daily and Dipirone 25mg/Kg by intravenous or intramuscular injection up to twice daily). Opioids were only used in the first two days after surgery (Morphine 30mg by intravenous injection, twice daily).

Antibiotics (Cefazoline 1g, by intravenous or intramuscular injection) were maintained for five days postoperatively.

Wound cleaning and dressing with dexamethasone, neomycin, nystatin, and benzocaine cream were performed twice daily.

### 4.9. Re-evaluation reintervention

After four weeks, the animals underwent a second procedure involving laparotomy and angiography. They were checked for vessel patency, patch infection or rupture, and leaks. The same anaesthetic and surgical protocols previously followed for the follow-up procedure.

Immediately after re-evaluation, while still under general anaesthesia, all animals were humanely euthanized with a Potassium Chloride solution. The aortic explant was performed for morphological analysis (Figure 5).

**Figure 5-.**
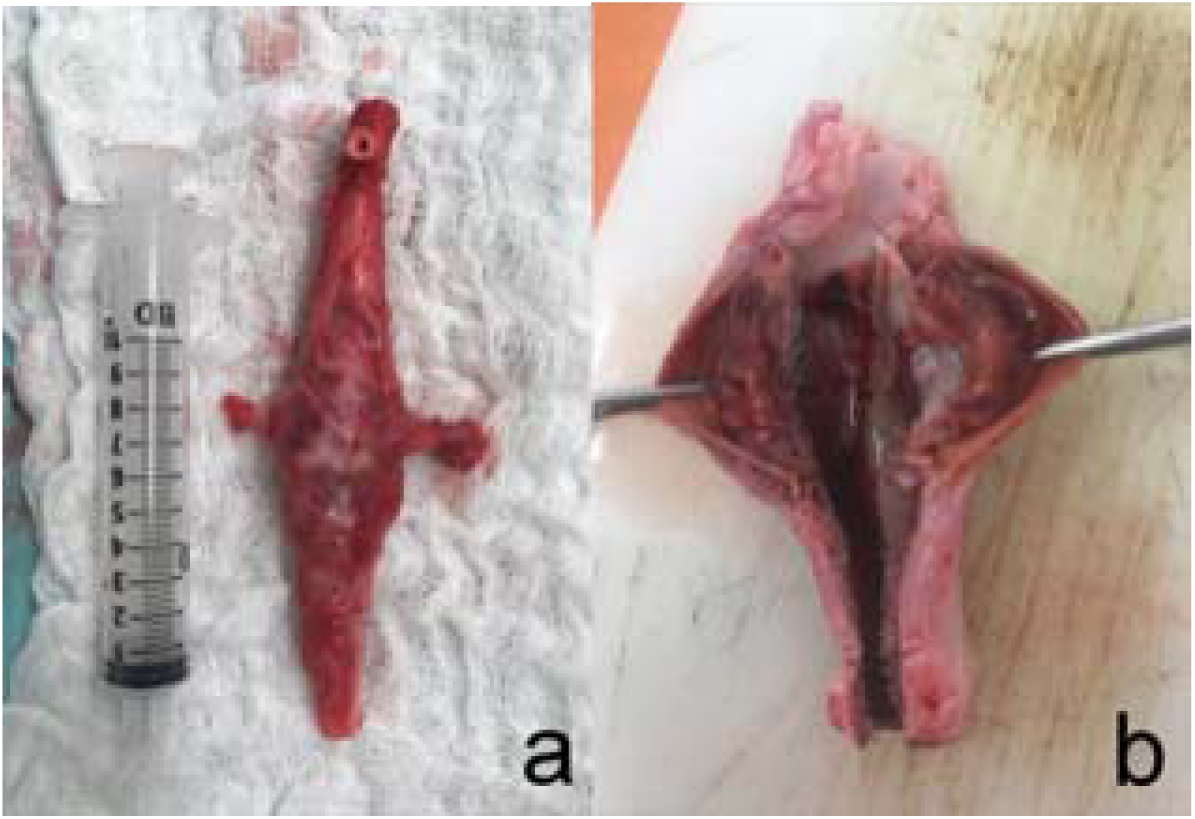
**A**: Aspect after explantation of the aneurysmal aorta; **B** Morphological aspect of the aneurysm sac.

Animals were observed for four weeks. The average weight gain during this interval was 12.06 ± 8.09 Kg.

## 5. Statistical Analysis

Categorical data were expressed as absolute frequencies and percentages, while continuous data were expressed as means with standard deviations and minimum-maximum values.

Associations between categorical data in the study groups were investigated using Fisher’s exact test. Analysis of continuous data was carried out using the Mann-Whitney test.

Linear models were adjusted to assess the effects of each technique application on each group. The results are presented as mean-adjusted values with standard errors and 95% confidence intervals. The *p*-values were obtained from multiple comparisons between measurements and group procedures.

All analyses were performed using SPSS Statistics for Windows version 19.0 (IBM Corp, Armonk, NY). A p-value less than 0.05 was considered significant.

## 6. Results

### 6.1. Primary procedure

In our sample, the primary procedure success rate for aneurysm creation was 100%, and the survival rate was 88.88%.

The average operating time was 176.22 ± 40.96 min (range 105 - 255 min).

When the technique of creating the aneurysmal sac from the native peritoneum was replaced by the use of bovine pericardium prepared at a back table, the mean operating time reduced to an average of 171.87 ± 38.24 min (range 105 – 170 min).

Although the change in the technique resulted in a notable reduction in operating time, the difference was not statistically significant (*p*=0,207).

#### 6.1.1. Outcomes according to clamping type

In the study, 18 animals were included. Three subjects underwent single clamping, while 15 underwent two-step sequential clamping *(Table I).* There were no significant differences in operating time (*p*=0.207) or total clamping time (*p*=0.276) between groups. However, there was a significant reduction in suprarenal clamping time (*p*=0.05), with an average reduction of over thirty minutes.

**Table I:**
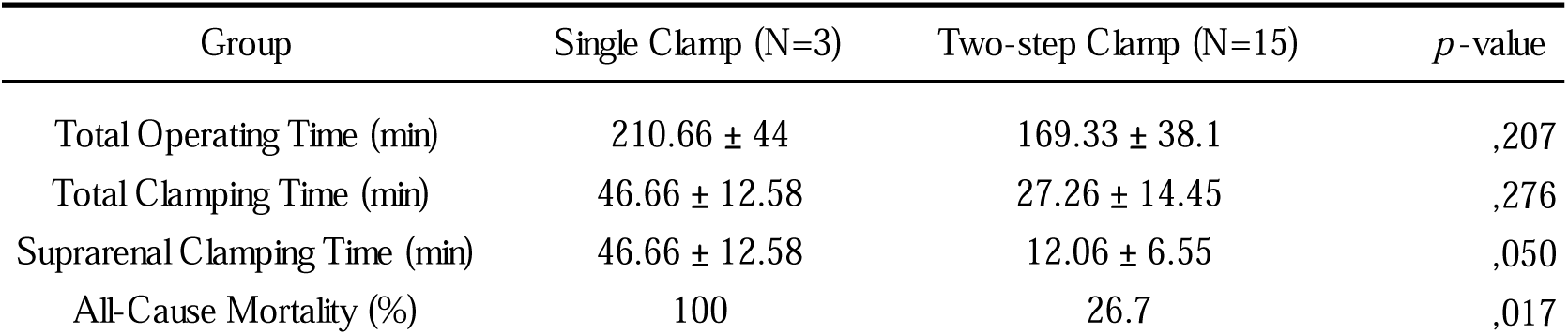
Single Clamp *versus* Two-step Sequential Clamp technique outcomes.

A significant association was found between death before 4 weeks and the single clamp technique (*p*=0.017), with the adoption of sequential clamping resulting in 73% reduction in death rates in this group. All animals (100%) in the single clamping group died, while in the sequential clamping group, only 4 (26.7%) animals died before the 4-week follow-up.

### 6.2. Follow-up and Study Protocol Completion

After four weeks, all of the surviving animals had patent aneurysms. There were no ruptures or infections of patches, and there were no occlusions in the aorta or its branches.

The completion rate of the animal protocol was 61.1%. Although 18 animals were initially enrolled, seven were lost during follow-up. The adverse outcomes of the animals are shown in Table II and discussed below. After euthanasia, aortic morphological analysis of all animals that did not complete the study showed the presence of patent aneurysms.

**Table II:**
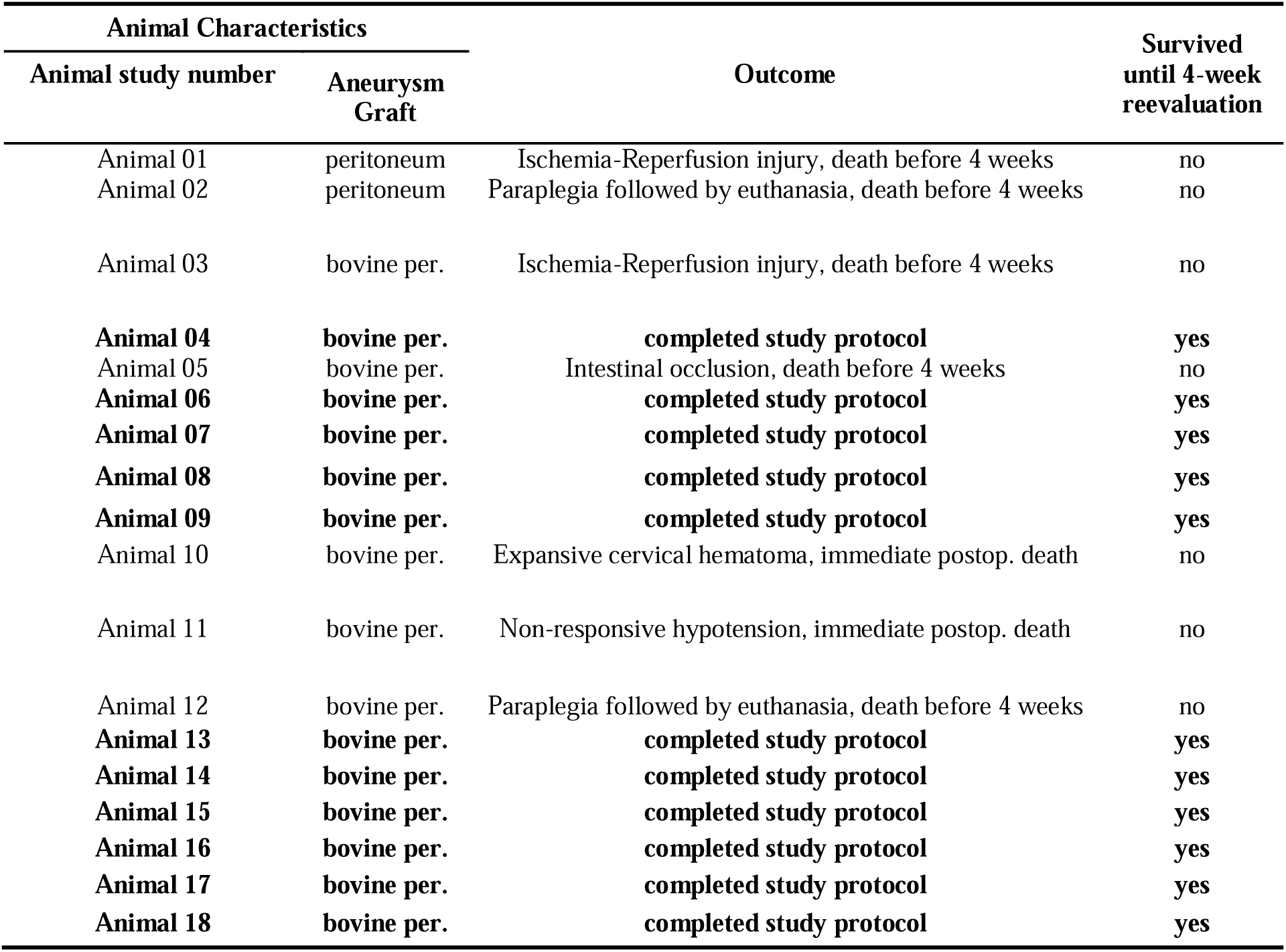
Individual animal outcomes.

## 7. Discussion

### 7.1. Animal Selection

It is nearly impossible to perfectly mimic the human physiopathological response, so there is no ideal experimental model. (11,12) Pigs, however, share similarities with humans regarding size, physiology, organ development, and disease progression, making them valuable models for human disease research. (13–15) Additionally, pigs have immune systems similar to humans, and inbred varieties allow for a fixed significant histocompatibility complex, granting reproducible results of immunologic mechanisms, unlike other models such as dogs and sheep. (16,17)

The large white breed has been validated as a good model for pharmacological trials, considering practical standpoints of size, weight, and development required for complying with various study protocols. (10)

In our sample, the animals weighed between 37 and 74 kg. We did not observe a maximum weight limit for this breed; however, the animal housing conditions in our centre had safety restrictions that were typically reached at around four weeks when the animals gained enough weight. Notably, the weight range for pig models in protocols of thoracoabdominal disease trials usually falls between 20 and 67 kg. (3,5,7,18,19)

The authors’ practice with swine experimental studies(13,14) has demonstrated that the 50 kg minimum weight threshold is ideal for surgical procedures due to the animal’s robust constitution and larger vessel diameter. However, these animals gain weight quickly, which could be a drawback for long-term follow-up.

### 7.2. Anaesthetic and preoperative procedures

In the scenario of experimental surgery in swine, it may be appropriate to administer preoperative medications to reduce anxiety and minimize the amount of general anaesthetic drugs needed.(20)

Veterinary literature also suggests that anticholinergics may help prevent the vagal reflex during endotracheal intubation and manipulation of the cardiovascular and pulmonary systems. (20)

Inhalation general anaesthesia is the preferred method for pigs in most protocols.(20) However, proper administration of these agents requires investment in drug administration equipment and monitoring physiological parameters.

Injectable anaesthetics are convenient for inducing anaesthesia before administering an inhalant agent or for short-term procedures(20).

### 7.3. Aneurysmal patch creation

The aneurysmal wall was originally constructed using a patch from the animal’s native peritoneum, as suggested by Maynar et al. (10) This technique could potentially eliminate the need for additional material while creating an autogenic aneurysmal sac. As a result, it may respond more like organic tissue and have the potential for further sac growth. (10)

The initial premise turned out to be flawed because using the peritoneum graft technique exponentially added to the operative time, incurring complications such as ischemia-reperfusion syndrome and hemodynamic instability. Maynar and his colleagues reported an average operative time of 120 minutes, with aortic occlusion time varying from 60 to 95 minutes (78 ± 16 minutes). However, in their study, only 4 out of 27 pigs subjected to aneurysm creation survived the 60-day follow-up. (10) The authors attributed the loss of animals to several complications, including acute renal failure, intestinal obstruction, pulmonary embolism, and two deaths from unknown causes. Additionally, two other subjects were euthanized as a consequence of paraplegia and extreme weight loss. The aneurysm rupture rate in the first two weeks of follow-up was 55.5%.(10)

In this study protocol, the peritoneum patch technique was discontinued after the second case in favour of the bovine pericardium patch. This approach resulted in a notable decrease in total operative time (176.22 ± 40,96 to 171.87 ± 38.24 min; *p*=0.207), although a significant correlation was not determined.

Furthermore, the previously described technique of sequential clamping significantly impacted the animals’ hemodynamic patterns and electrolyte balance during both the surgical and postoperative periods. The mortality rate among animals in the Sequential Clamping group (26.7%) was much lower than that of the Single Clamping group (100%). The high mortality rate in the Single Clamping group is consistent with findings from another animal aortic experimental studies.

The technique of creating an artificial patch in a saccular shape, proposed by Uflacker and Brothers,(21) does not offer the advantage of mimicking the shape and size configurations of the conventional TAAA, as would, for example, the patch interposition model suggested by Parodi *et al* (22). Unlike the Parodi configuration, the saccular approach provides more accurate results regarding the role of the biologic aneurysmal wall and the effects of the patent aortic side branches, which were indispensable when analysing the thoracoabdominal territory. (22)

Several studies have effectively used cow pericardium patches to create artificial aneurysms in experimental models, achieving good patency rates. (7,23,24) These patches have the ability to expand, simulating an aneurysmal segment due to antigenic degeneration. However, because it is an acellular material, this response is minimal and does not incur high rupture rates like vein grafts or their other synthetic counterparts. (25) The literature describes patch sizes for aneurysm creation ranging from 3 cm x 3 cm to 3 cm x 6 cm in pigs and 6 cm x 5 cm x 8 cm in sheep. (23)

### 7.4. Postoperative analgesia and euthanasia

After surgery, pigs may show signs of distress, such as restlessness and refusal of food. (20) Newer non-steroidal anti-inflammatory drugs (NSAIDs) have been utilized successfully for postoperative pain relief, given either orally or by injection. Parenteral analgesics are usually administered intramuscularly or subcutaneously in the neck. Pigs can be easily encouraged to take oral medication by mixing it with their food. (20)

For euthanasia, most injectable forms used in other large animal species are suitable for pigs. Pentobarbital overdose (>150 mg/kg) is the preferred form of parenteral euthanasia. (20) Potassium Chloride injections or exsanguination may be performed while the pig is under general anaesthesia. (26)

### 7.5. Protocol Outcomes

**Primary Procedures:** In our experimental model, we achieved a 100% success rate in creating aneurysms and had no deaths during the primary procedure. In most experimental swine models, primary procedures had high success rates and few complications. In a report by Bischoff *et al*., pigs were subjected to deployment of an endograft in the abdominal aorta to evaluate spinal ischemia, with the authors describing only one complication: one stent migration covering the celiac axis.(19)

Okuno and colleagues designed an experimental model using pigs to evaluate thoracoabdominal dissections. In the primary procedure, an artificial aortic dissection was created with a 78.6% success rate. The three failed cases described include one guide wire trapping in the false lumen and two intraoperative aortic ruptures.(3)

Two experimental swine trials previously published by our group, either to evaluate aorta stent-graft oversizing (13) or to assess the rheological effect of renal ischemia and reperfusion in pigs, also showed a 100% success rate and a 100% survival rate intraoperatively. (8)

Finally, the technical success and survival rates in our triaĺs primary procedure do not remarkably differ from those reported by authors who performed similar artificial aortic saccular aneurysm creations. In the reports by Kalder *et al. and* Aquino *et al., the* technical success of aneurysm creation was 100% also (7,23).

### 7.6. Follow-up and protocol completion

In our trials, follow-up intervals are mostly limited due to the limited space for animals in our Centre for Experimentation and Training in Surgery, particularly for the large white pig breed, which gains weight relatively quickly in confinement. (8,19) Monitoring these animals for an extended period would compromise their well-being due to their size and weight. Therefore, we have found that a four-week follow-up period is typically sufficient for the animals to develop enough for outcomes to be perceived without jeopardizing their welfare.

The bovine pericardium patches showed 100% aneurysm patency after weeks. Similarly designed trials also reported 100% aneurysm patency after 2 weeks,(7) and at the 52-week follow-up.(23) Aquino described parietal thrombus formation in all subjects, with two occluded (18%; IC 95% = 3.98 – 48.84) and nine patent aneurysms (82%; IC 95% = 51.15 – 96.01). (7)

In our sample, no ruptures nor aortic or branch occlusions were encountered. In the trial conducted by Kalder and collaborators, they reported one case of haemorrhage from the aneurysmal suture line and one case of infection and rupture of the aneurysmal patch. (23)

The animal protocol completion rate in our trial was 61.1%. Okuno *et al.* conducted follow-up observations in five of the original 14 animals. In this sample of five pigs, two died before the 5-day survey follow-up due to confirmed aortic rupture. (3) In Kalder’s report, 2 of the 6 original subjects died before protocol completion. The first animal died within one week as a consequence of aneurysmal suture line bleeding, and the second one died after 48 weeks due to infection and rupture of the aneurysmal patch. (23) Aquino, however, reports protocol completion of all eleven animals in his sample. (7)

Our study found that the main reasons for animal loss during follow-up were long intervals of aortic clamping. According to Table II, 7 animals were lost in surveillance, with 5 perishing due to consequences of increased clamping time. Ischemia and reperfusion injury (N=3) or paraplegia induced by spinal ischemia (N = 2) were the main causes of animal loss. Additionally, there were cases of digestive tract complications and respiratory insufficiency following cervical expansive hematoma.

In the “Impact of Stent-Graft Oversizing on the Thoracic Aorta: Experimental Study in a Porcine Model” trial, the iliac arteries were accessed through retroperitoneal access, (13) avoiding the need for peritoneal exposure. However, for the implantation of the thoracoabdominal aneurysmal patch and retraction of both renal arteries, the aorta had to be exposed at the level of the visceral branches, which was hindered laboriously by this approach. As a result, pigs were, therefore, subjected to transperitoneal access with aortic clamping. Although a direct comparison of these two samples has not been conducted, it was noted that the late mortality rate was remarkably higher in this latter trial (0% *versus* 44,4%), as is commonly seen in procedures that involve peritoneal exposure and suprarenal clamping.

### 7.7. Pitfalls

While developing the protocols for our trials, we learned that a shorter operative time is paramount for minimizing postoperative complications and animal loss. We shortened the operative time by adopting a back table system to create an aneurysmal patch from the bovine pericardium, with two parallel teams working together.

Carefully planned anaesthetic procedures and establishing a proper intensive postoperative care system for the swine models are essential for minimizing the high mortality inherent in procedures of this stature.

Several factors must be considered when applying an experimental animal model. No animal model will ever perfectly mimic human pathophysiology, so it is important to use multiple models to gain a better understanding before choosing the most appropriate one. This means it is crucial to clearly define the hypothesis to develop an experimental protocol that could lead to relevant clinical data.

## 8. Conclusion

In the presented swine model, to use a bovine pericardium patch reduce the duration of the operation. The use of a sequential two-step clamping significantly decreases the duration of suprarenal clamping time and overall mortality.

The experimental model of a thoracoabdominal bovine pericardium aneurysm which uses a sequential clamping technique, provides a stable and reliable platform with consistent anatomy and patency for up to four weeks. This model can be extremely valuable for assessing new endovascular therapy options in living organisms.

## Data Availability

All data produced in the present work are contained in the manuscript

